# Impact of an Early Body Mass Index Maintenance Intervention at ART initiation on Diabetes Risk: Insights from a Simulation Study

**DOI:** 10.1101/2025.11.05.25339607

**Authors:** Parastu Kasaie, Yao Zhao, Elizabeth Humes, Lucas Gerace, Kendall Reid, Catherine R. Lesko, Katherine E. Kurgansky, John R. Koethe, Peter F. Rebeiro, Kassem Bourgi, Kristine M. Erlandson, Kelly Gebo, Keri N Althoff

## Abstract

**Background:** Weight gain after antiretroviral therapy (ART) initiation is linked to increased risk of developing type 2 diabetes mellitus among people with HIV (PWH). This study examines the potential impact of a hypothetical body mass index (BMI) maintenance intervention after ART initiation on reducing diabetes risk among PWH in the United States (US).

**Methods:** Using an agent-based simulation of comorbidities in PWH initiating ART in the US, we simulated a hypothetical two-year intervention, where BMI remained stable for individuals who would otherwise gain BMI. Individuals starting ART between 2013–2017 with a normal or overweight BMI and no prior diabetes diagnosis were eligible to participate. Participants were followed under an intervention scenario and a control scenario with no intervention. Diabetes risk was calculated as new diagnoses per 1,000 person-years over seven years after ART initiation.

**Results:** The model forecasted that 171,000 PWH would initiate ART between 2013–2017, with 73% meeting eligibility criteria. Without intervention, 66% were expected to gain BMI, and 12% would become obese. The forecasted diabetes risk was 10.7 per 1,000 person-years, with approximately 9,500 new diagnoses over seven years after ART initiation. Under the hypothetical intervention, diabetes risk was reduced by 19% and a median of 1,800 diagnoses were averted. Subgroup analyses showed the largest risk reductions among women who inject drugs and Hispanic heterosexual men.

**Conclusion:** Preventing BMI increases in ART initiators could significantly reduce diabetes risk and improve health outcomes among PWH. Tailored weight maintenance strategies are needed to address diverse subgroup needs.

## INTRODUCTION

The landscape of HIV epidemiology in the United States (US) has changed significantly since the advent of antiretroviral therapy (ART). Prior to ART, wasting was common, and after ART introduction, weight gain after treatment initiation was seen as a “return to health” phenomenon accompanying immunologic and nutritional recovery. Today, the majority of people with HIV (PWH) initiate ART with a CD4 count >300 cells/mm^3^,^1^ and an increasing proportion are overweight or obese at ART initiation.^2^ In the North America AIDS Cohort Collaboration on Research and Design (NA-ACCORD), the proportion of individuals with obesity at ART initiation in the US and Canada increased from 11% (1998–2000) to 17% (2007–2010)^3^, reflecting broader trends.^4-6^ Many PWH experience excess weight gain after ART, with an average annual Body Mass Index (BMI) increase of 0.53 (kg/m^2^) for PWH versus 0.12 (kg/m^2^) for matched individuals without HIV.^7^ While weight gain is beneficial for underweight individuals, it often results in excess fat for those already overweight.^8-13^ After three years of follow up, 22% of normal-weight individuals at initiation became overweight, while 18% of overweight progressed to obesity.^3^ Black/African Americans (Black), women, those with low pre-ART CD4 count and higher HIV-1 RNA are more likely to gain weight on ART.^3,8,14-18^ Some common antiretrovirals including integrase strand transfer inhibitors (INSTIs) and tenofovir alafenamide (TAF) have also been independently linked to greater weight gain.^14,16,19,20^

With prolonged survival on ART and rising comorbidities among PWH in the US, addressing weight gain has become crucial for both lifespan and the health span. Obesity in PWH is strongly linked to metabolic diseases such as type 2 diabetes.^21-23^ Preventing BMI increases in normal or overweight individuals during the initial years of ART could significantly reduce diabetes risk. While no clinical trials have specifically studied weight maintenance at ART initiation and its long-term effects on diabetes risk, advanced computational techniques can offer insights into its potential impact. Using a computer simulation of PWH receiving ART in the US, we evaluated the impact of preventing BMI increases during the first 24 months after ART initiation on diabetes risk over the first seven years on treatment.

## METHODS

ProjEcting Age, MultimoRbidity, and PoLypharmacy (PEARL) is an agent-based simulation of PWH initiating ART in the US (**Figure 1**).^24-27^ The model leverages the NA-ACCORD data from 2009-2017 (**Supplement S1**) to inform projections among 15 sub-groups of PWH including men who have sex with men (MSM), women who inject drugs (WWID), men who inject drugs (MWID), and heterosexual men and women, from Hispanic, non-Hispanic White, or non-Hispanic Black populations.^28^

**Figure 1:**
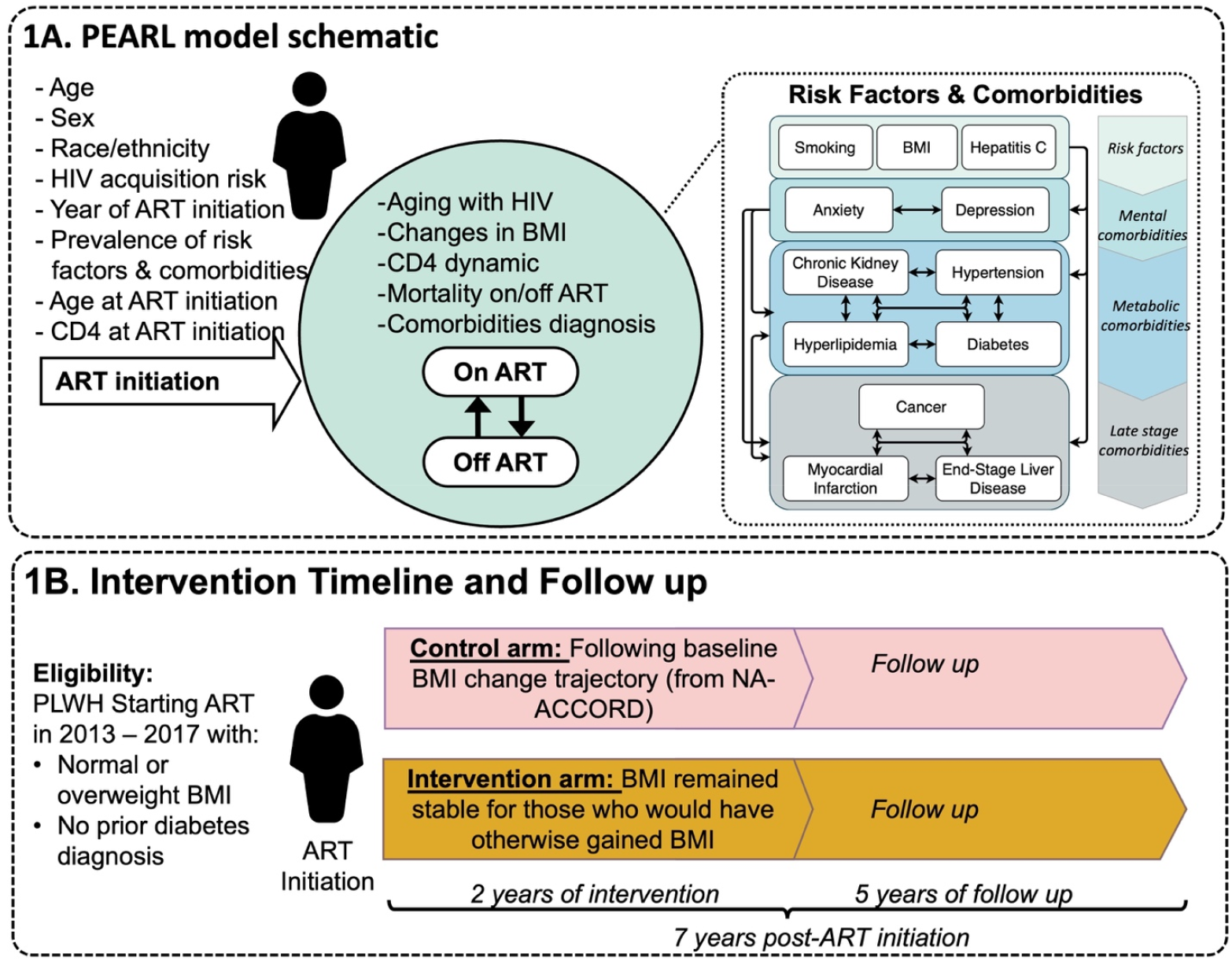
Model schematic of PEARL simulation. **Panel A** shows a schematic figure of simulated agents in the PEARL model. Agents enter at ART initiation and are followed to the end of simulations in 2035. The model includes the dynamics of aging, ART engagement and disengagement, changes in CD4 count, and mortality risk in each calendar year. Changes in BMI are modeled from ART initiation to 24 months after ART. Mathematical functions simulate the prevalence of risk factors and comorbidities at ART initiation and changes in these over time via the probability of incident events, reflecting the medical complexity that can occur as people age with HIV. **Panel B** illustrates the drivers of diabetes diagnosis risk in the PEARL model. The risk (probability) of incident diabetes is estimated from a logistic regression model that incorporates these key individual-level drivers. **Panel C** illustrates the logic for the simulated clinical trial focused on a “hypothetical” BMI maintenance intervention after ART initiation. PWH starting ART between 2013-2017 with normal or overweight BMI and no pre-existing diabetes diagnosis are eligible to participate in the study. Simulated study includes (1) a control arm, where agents experienced no intervention affecting their BMI after ART initiation or subsequent diabetes risk, and (2) an intervention arm, where eligible agents participated in a two-year BMI maintenance program. Individuals in both arms are monitored for five years post intervention (follow-up continuing through 2024 for agents who initiated ART in 2017) to record incident diabetes diagnoses over seven years after ART initiation. Abbreviations: ART: Antiretroviral treatment, BMI: Body Mass Index, PWH: People with HIV

Briefly, PEARL follows the life course of PWH after ART initiation, starting in 2009 with an initial cohort of PWH who initiated ART in the US. Each year, a new cohort of ART initiators is added (**Supplement S2.1**). Simulated individuals experience annual risk of ART disengagement, ART reengagement, mortality, and diagnoses with age-related comorbidities including mental (depression, anxiety), metabolic (dyslipidemia, hypertension, diabetes, chronic kidney disease) and late-stage diseases (cancer, myocardial infarction, end stage liver disease). Additionally, the model incorporates three risk factors influencing comorbidity risk: cigarette smoking, hepatitis C virus infection, and BMI categories. These probabilities are estimated using regression models informed by NA-ACCORD data (**Supplement S2.2 & S2.3**). Further details on the parametrization and validation of PEARL’s forecasts can be found in previous publications^24-27^ and on the PEARL website: https://pearlhivmodel.org.

### Simulated BMI Maintenance Intervention at ART Initiation

We hypothesize that for groups starting ART with normal or overweight BMI, increases in BMI during the first 24 months after ART initiation are linked to a heightened risk of developing diabetes.^29-31^ To this end, we simulated a “hypothetical” BMI maintenance intervention for PWH starting ART with a normal or overweight BMI (18.5-30 kg/m^2^) and without a prior diabetes diagnosis. The intervention, lasting two years, allowed BMI to decrease naturally but restricted any increases, maintaining BMI at ART initiation levels for individuals who would otherwise have gained BMI.

We modeled the intervention’s impact under two scenarios: (1) a control arm, where agents experienced no intervention affecting their BMI or subsequent diabetes risk, and (2) an intervention arm, where all eligible agents participated in the two-year BMI maintenance program.

NA-ACCORD has limited data on populations using INSTI-based regimens by 2017. To accommodate this restriction, we excluded ART regimen from our analysis, and focused on the 2013–2017 period, when a mix of ART regimens was used, minimizing potential impact on future projections. The two-year intervention was simulated over five years (2013-2017), and individuals were monitored for changes in diabetes risk over five years following the program (total of seven years after ART initiation, **Figure 1**). The primary analysis assumes full program coverage (i.e., 100% of eligible agents were enrolled) and complete adherence (i.e., the program was 100% effective in maintaining agents’ BMIs).

### Primary Outcomes

The risk of incident diabetes diagnosis over the seven-year following ART initiation was estimated by dividing the total number of new diabetes diagnoses by the total number of person-years at risk. Simulated individuals were censored at death. We report both the absolute and relative reductions in diabetes risk between the simulated intervention and control scenarios. Additionally, we estimate the number needed to treat (NNT) to prevent one diabetes diagnosis for each subgroup. To assess the population-level impact, we report the total number of diabetes diagnoses averted in each subgroup over seven-years of follow-up. A total of 1,000 simulations were performed across all subgroups and scenarios to obtain stable estimates for the primary outcomes.

### Sensitivity Analysis

We conducted a probabilistic sensitivity analysis to evaluate variations in six primary parameters including the number of ART initiators, BMI at- and after-ART initiation, diabetes prevalence among initial ART users and ART initiators, and incidence diabetes diagnosis among ART users. Each parameter was randomly perturbed within ±20% of its baseline value. Sensitivity was assessed by comparing simulations using the parameters in their highest and lowest decile. Furthermore, we assessed the sensitivity of primary outcomes to reductions in intervention coverage and effectiveness, adjusting both to 75% within each subgroup.

## RESULTS

### Simulated Population Characteristics

PEARL model forecasted a median of 171,000 [95% Uncertainty Range (UR): 169,000–173,000] PWH initiating ART in the US from 2013-2017 (**Table 1**). Black MSM and Hispanic WWID were the youngest and oldest subgroups at ART initiation, with median ages of 29 [21–54] and 48 [27–65] years, respectively. The model forecasted that 5% of those initiating ART had a pre-existing diabetes diagnosis, with the lowest prevalence among White and Hispanic MSM (3%), and the highest prevalence among Black MWID (13%). At ART initiation, 3% of ART initiators were underweight (BMI<18.5 kg/m^2^), and 19% were obese (BMI≥30 kg/m^2^). Obesity rates were higher among women, particularly Black heterosexual women (41%) and Black WWID (33%). Among men, MSM subgroups had the lowest obesity proportion (13%) at ART initiation. After excluding individuals with diabetes, underweight, or obesity at ART initiation, 73% (126,000 [124,000–127,000]) of ART initiators between 2013–2017 met the eligibility criteria for the simulated BMI maintenance intervention.

**Table 1:** Population characteristics of simulated agents starting ART from 2013–2017 in the US. Values represent median [95% uncertainty range] across 1,000 random simulations at baseline (without additional interventions). Population size numbers are rounded to the nearest hundred and percentages are rounded to zero.

| Group | Population Size of ART initiators | Age at ART initiation | Population percentage with pre-existing diabetes diagnosis at ART initiation | Population percentage with BMI <18.5 at ART initiation | Population percentage with 18.5≤ BMI <30 at ART initiation | Population percentage with BMI ≥30 at ART initiation | Population percentage of ART initiators eligible for the intervention | Number of ART initiators eligible for the intervention | Percentage of eligible population experiencing weight gain over 2-years following ART initiation | Percentage of eligible population developing obese BMI (≥30) over 2-years following ART initiation |
| --- | --- | --- | --- | --- | --- | --- | --- | --- | --- | --- |
| **Black HET Women** | 19,400<br>[18,800–20,100] | 41<br>[24–59] | 10%<br>[9–10%] | 2%<br>[2–2%] | 56%<br>[55–56%] | 41%<br>[40–41%] | 50%<br>[49–51%] | 9,900<br>[9,500–10,200] | 71%<br>[70–72%] | 24%<br>[24–25%] |
| **White HET Women** | 4,400<br>[4,200–4,500] | 41<br>[25–62] | 10%<br>[9–10%] | 4%<br>[4–5%] | 64%<br>[63–65%] | 30%<br>[29–31%] | 58%<br>[57–59%] | 2,600<br>[2,500–2,700] | 63%<br>[61–64%] | 16%<br>[15–17%] |
| **Hispanic HET Women** | 5,100<br>[4,900–5,300] | 38<br>[25–65] | 10%<br>[9–10%] | 1%<br>[1–2%] | 62%<br>[61–63%] | 35%<br>[34–36%] | 56%<br>[55–57%] | 2,900<br>[2,800–3,000] | 67%<br>[65–68%] | 18%<br>[17–19%] |
| **All HET Women** | 28,800<br>[28,100–29,500] | 41<br>[24–61] | 10%<br>[9–10%] | 2%<br>[2–2%] | 58%<br>[58–59%] | 38%<br>[38–39%] | 52%<br>[52–53%] | 15,200<br>[14,800–15,600] | 69%<br>[68–69%] | 22%<br>[21–22%] |
| **Black HET Men** | 9,400<br>[9,100–9,700] | 43<br>[24–61] | 9%<br>[8–9%] | 3%<br>[3–3%] | 74%<br>[74–75%] | 21%<br>[20–22%] | 67%<br>[66–68%] | 6,400<br>[6,200–6,600] | 68%<br>[67–69%] | 14%<br>[13–15%] |
| **White HET Men** | 2,100<br>[2,000–2,200] | 46<br>[26–66] | 9%<br>[8–10%] | 2%<br>[2–3%] | 80%<br>[78–81%] | 16%<br>[15–18%] | 72%<br>[71–74%] | 1,500<br>[1,400–1,600] | 75%<br>[73–77%] | 15%<br>[14–17%] |
| **Hispanic HET Men** | 2,900<br>[2,800–3,000] | 43<br>[26–67] | 9%<br>[8–10%] | 2%<br>[2–3%] | 80%<br>[79–81%] | 16%<br>[15–17%] | 72%<br>[71–74%] | 2,100<br>[2,000–2,200] | 79%<br>[78–81%] | 20%<br>[19–22%] |
| **All HET Men** | 14,300<br>[13,900–14,700] | 44<br>[24–63] | 9%<br>[9–9%] | 3%<br>[3–3%] | 76%<br>[76–77%] | 19%<br>[19–20%] | 69%<br>[68–70%] | 10,000<br>[9,700–10,200] | 71%<br>[71–72%] | 16%<br>[15–16%] |
| **Black WWID** | 2,000<br>[1,800–2,300] | 48<br>[27–65] | 10%<br>[8–11%] | 2%<br>[2–3%] | 63%<br>[62–65%] | 33%<br>[31–34%] | 57%<br>[55–59%] | 1,200<br>[998–1,300] | 60%<br>[57–62%] | 17%<br>[15–18%] |
| **White WWID** | 2,100<br>[2,000–2,300] | 41<br>[26–60] | 10%<br>[8–11%] | 3%<br>[2–3%] | 76%<br>[74–77%] | 20%<br>[19–22%] | 68%<br>[66–70%] | 1,500<br>[1,400–1,600] | 76%<br>[74–78%] | 24%<br>[22–25%] |
| **Hispanic WWID** | 863<br>[792–932] | 47<br>[31–62] | 10%<br>[8–11%] | 3%<br>[2–4%] | 77%<br>[75–79%] | 18%<br>[16–21%] | 69%<br>[67–72%] | 601<br>[548–656] | 73%<br>[70–76%] | 20%<br>[18–23%] |
| **All WWID** | 5,000<br>[4,700–5,300] | 45<br>[27–63] | 10%<br>[9–10%] | 3%<br>[2–3%] | 71%<br>[70–72%] | 25%<br>[24–26%] | 64%<br>[63–65%] | 3,200<br>[3,000–3,400] | 69%<br>[68–71%] | 21%<br>[19–22%] |
| **Black MWID** | 3,900<br>[3,500–4,400] | 45<br>[22–63] | 13%<br>[13–14%] | 2%<br>[2–3%] | 79%<br>[78–80%] | 17%<br>[16–18%] | 68%<br>[67–69%] | 2,700<br>[2,400–3,000] | 58%<br>[56–59%] | 8%<br>[7–9%] |
| **White MWID** | 5,600<br>[5,300–5,900] | 43<br>[24–62] | 6%<br>[5–6%] | 2%<br>[2–3%] | 88%<br>[87–89%] | 8%<br>[7–9%] | 82%<br>[82–83%] | 4,600<br>[4,400–4,900] | 63%<br>[62–64%] | 8%<br>[7–9%] |
| **Hispanic MWID** | 3,500<br>[3,300–3,700] | 42<br>[24–59] | 6%<br>[5–6%] | 0%<br>[0–0%] | 70%<br>[69–72%] | 28%<br>[27–29%] | 66%<br>[65–67%] | 2,300<br>[2,200–2,500] | 57%<br>[55–59%] | 11%<br>[10–12%] |
| **All MWID** | 12,900<br>[12,300–13,600] | 43<br>[23–62] | 8%<br>[8–9%] | 2%<br>[2–2%] | 81%<br>[80–81%] | 16%<br>[16–17%] | 74%<br>[73–74%] | 9,600<br>[9,100–10,100] | 60%<br>[59–61%] | 9%<br>[8–9%] |
| **Black MSM** | 43,400<br>[42,400–44,400] | 29<br>[21–54] | 4%<br>[4–4%] | 4%<br>[4–4%] | 80%<br>[79–80%] | 15%<br>[15–15%] | 76%<br>[76–77%] | 33,400<br>[32,600–34,100] | 67%<br>[67–67%] | 11%<br>[11–11%] |
| **White MSM** | 33,200 | 40 | 3% | 2% | 84% | 12% | 81% | 27,200 | 66% | 9% |
|  | [32,700–33,800] | [23–60] | [3–3%] | [2–2%] | [84–85%] | [12–13%] | [81–82%] | [26,700–27,700] | [65–66%] | [8–9%] |
| **Hispanic MSM** | 33,300<br>[32,300–34,500] | 32<br>[22–52] | 3%<br>[3–3%] | 2%<br>[2–2%] | 85%<br>[84–85%] | 12%<br>[12–13%] | 82%<br>[81–82%] | 27,400<br>[26,600–28,400] | 65%<br>[65–66%] | 8%<br>[8–9%] |
| **All MSM** | 109,900<br>[108,200–111,600] | 32<br>[21–56] | 3%<br>[3–3%] | 3%<br>[3–3%] | 83%<br>[82–83%] | 13%<br>[13–13%] | 79%<br>[79–80%] | 87,900<br>[86,500–89,300] | 66%<br>[66–66%] | 9%<br>[9–10%] |
| **Overall** | 170,800<br>[168,700–172,800] | 36<br>[22–59] | 5%<br>[5–5%] | 3%<br>[2–3%] | 77%<br>[77–78%] | 19%<br>[18–19%] | 73%<br>[73–73%] | 125,700<br>[124,200–127,200] | 66%<br>[66–67%] | 12%<br>[11–12%] |

Abbreviations: ART: Antiretroviral treatment, BMI: Body Mass Index, Black: Black/African American, Het: Heterosexual, WWID: Women Who Inject Drugs, MWID: Men Who inject Drugs, MSM: Men who have Sex with Men

### Change in BMI After ART Initiation and the Risk of Incident Diabetes Diagnosis in the Control Arm

In the absence of the intervention, 66% of ART initiators in the control arm experienced a BMI increase (median increase 1.01 kg/m^2^ [0.39–2.38]), with 12% becoming obese over the 2 years. Subgroup analysis revealed significant heterogeneities in BMI trajectories, with Hispanic heterosexual men experiencing the highest risk of BMI gain (79%) and reaching obesity (24%). Among the persons who inject drug, White and Hispanic WWID gained the most weight, with over 70% gaining BMI and over 20% becoming obese, while MWID had lower rates, with 60% gaining BMI and 9% becoming obese.

In the seven years following ART initiation, a median of 9,460 participants [9,260–9,650] developed incident diabetes diagnosis, corresponding to a risk of 10.7 [10.6–10.9] per 1,000 person-years. This risk increased with age and BMI at ART initiation (**Figure 2A-B**). WWID experienced the highest risk, ranging from 26.4 to 29.9 per 1,000 person-years, followed by heterosexual women, ranging from 21.4 to 24.2 per 1,000 person-years across race and ethnicity subgroups (**Figure 2C**). Lowest risk was forecasted among Hispanic and White MSM at 3.4 and 4.5 per 1,000 person-years respectively. The largest number of incident diabetes diagnoses were forecasted among Black MSM (2,860 [2,760–2,980]), followed by Black heterosexual women (1,650 [1,570–1,750], **Figure 2D**).

**Figure 2.**
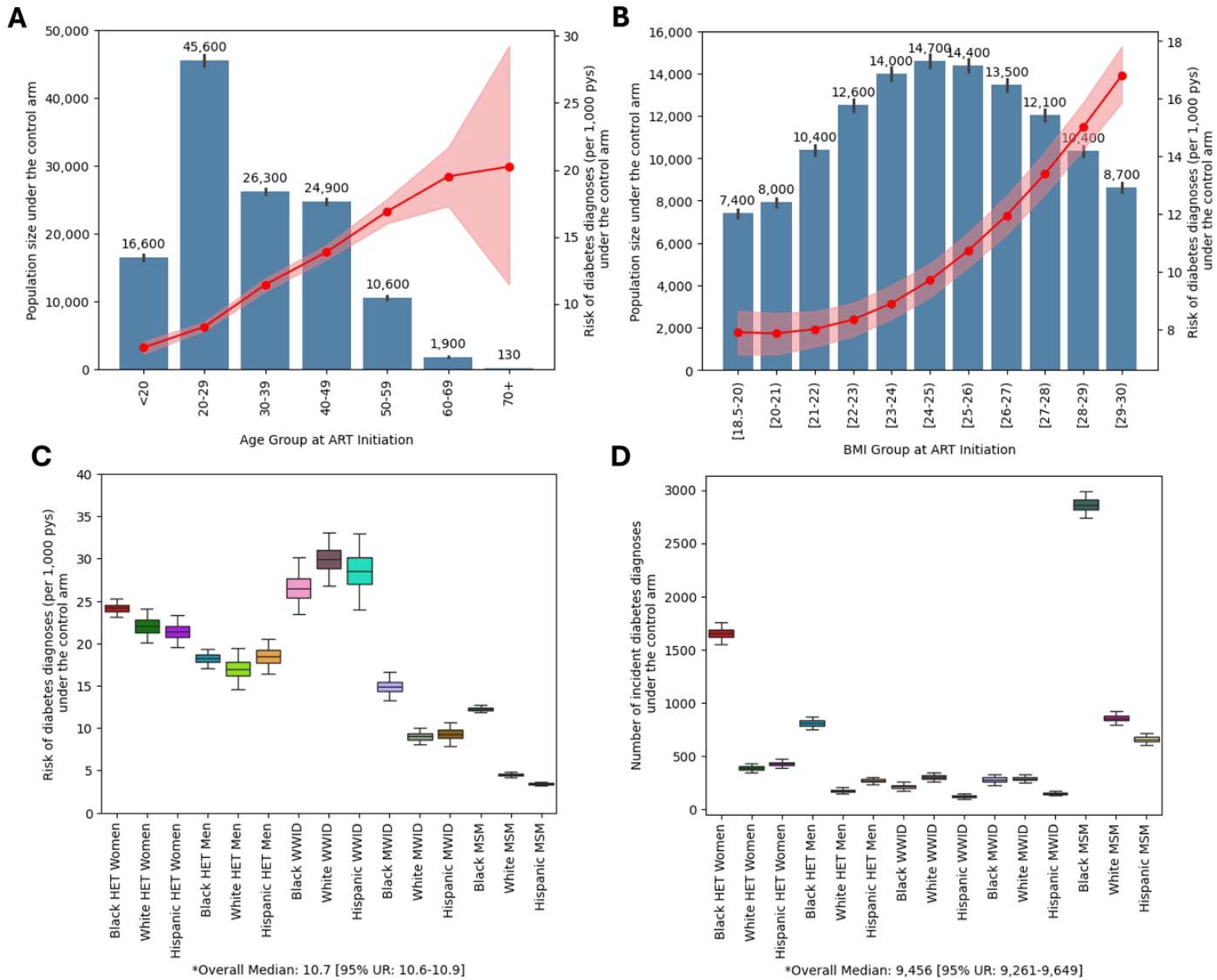
Forecasted risk of incident diabetes in the control arm. This figure illustrates the forecasted seven-year risk of incident diabetes among study participants under the control arm. The population consists of 15 subgroups of PWH starting ART between 2013–2017, with no prior diabetes diagnosis and a normal or overweight BMI at ART initiation. Participants were enrolled in a two-year placebo intervention and followed for an additional five years to assess the risk of diabetes diagnosis. Results are derived from 1,000 random simulations under each arm. **Panel A** shows the distribution of age at ART initiation (primary y axis) with bars indicating median values (shown above each bar) and whiskers representing the 95% uncertainty range. The red line represents the forecasted median seven-year risk of diabetes (per 1,000 person years at risk, secondary y axis) and the shaded area represents the 95% uncertainty range. **Panel B** shows the distribution of BMI at ART initiation with bars indicating median values (shown above each bar) and whiskers representing the 95% uncertainty range. Similarly, the red line and shaded area represent the forecasted median and 95% uncertainty range in seven-year risk of diabetes (per 1,000 person years at risk, secondary y axis). **Panel C** shows the distribution of the seven-year risk of incident diabetes diagnoses (the median and interquartile range represented in the box plots, with whiskers extending to 95 percentile range). **Panel D** shows the number of incident diabetes diagnoses (the median and interquartile range represented in the box plots, with whiskers extending to 95 percentile range). Abbreviations: PWH: people with HIV, ART: Antiretroviral treatment, BMI: Body Mass Index, Black: Black/African American, Het: Heterosexual, WWID: Women Who Inject Drugs, MWID: Men Who inject Drugs, MSM: Men who have Sex with Men

### Impact of a BMI Maintenance Intervention at ART Initiation on the Risk of Incident Diabetes Diagnosis

Among ART initiators receiving the intervention, the risk of incident diabetes diagnosis over the seven years following ART initiation was forecasted a median of 8.7 [8.5–8.9] per 1,000 person-years. This reflects an absolute reduction of 2.0 [1.7–2.3] diabetes diagnosis per 1,000 person-years compared to the control group (**Figure 3A**), translating to a relative reduction of 19% [16–21%] in risk (**Figure 3B**). At a population level, this intervention was forecasted to avert 1,760 [1,470–2,070] new diabetes diagnoses in the seven years after ART initiation, among 126,000 individuals receiving it (**Figure 3D**). This corresponds to 71 [61–85] ART initiators receiving the intervention to avert one diabetes diagnosis (**Figure 3C**).

**Figure 3.**
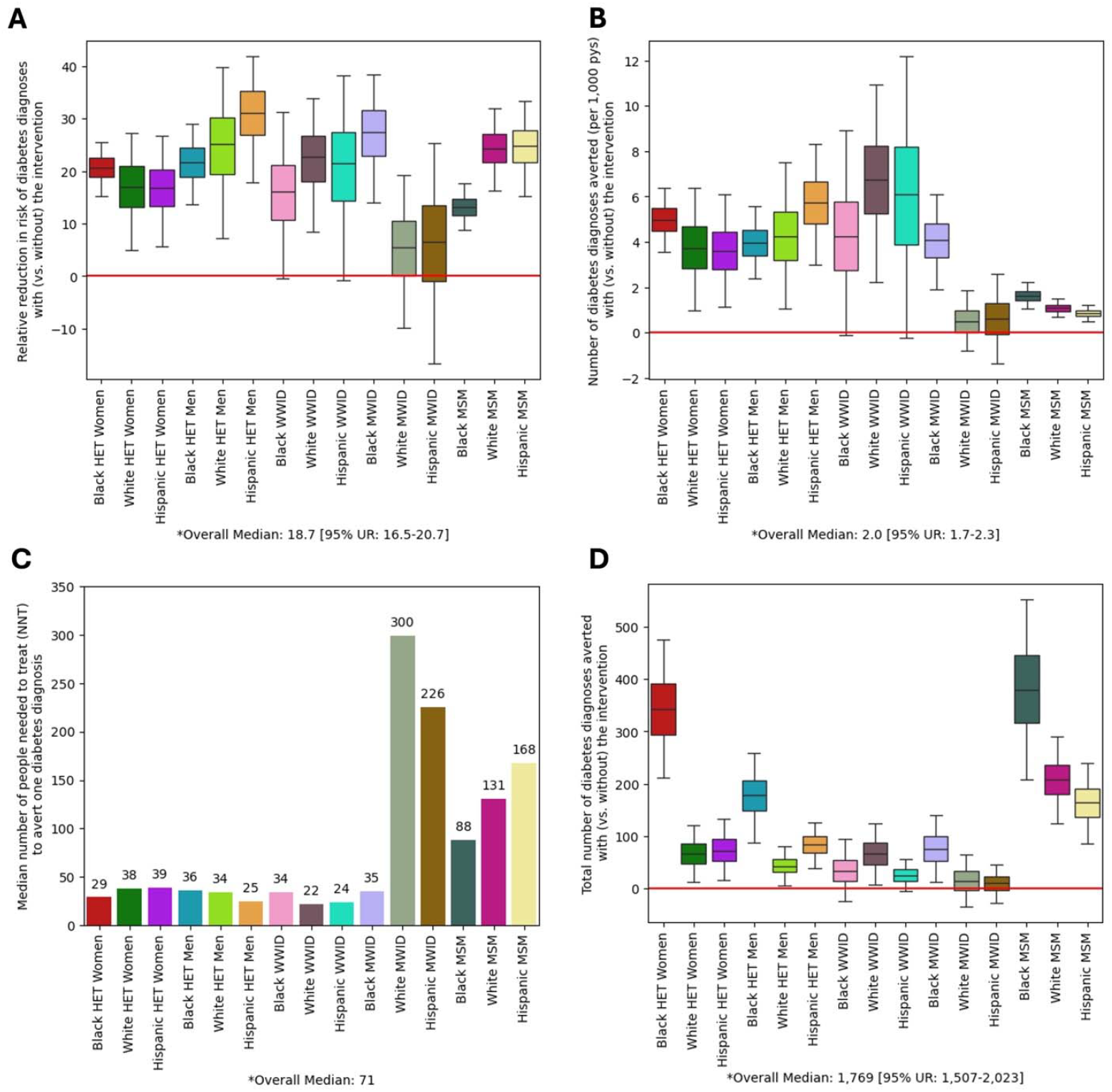
Forecasted impact of a BMI maintenance intervention at ART initiation on the risk of incident diabetes diagnosis. Simulated individuals in the intervention arm participate in a two-year BMI management program, where BMI remained stable for individuals who would have otherwise gained weight. In contrast, individuals in the control arm follow the expected trajectory of weight change after ART initiation. The primary outcome of interest is the difference in risk of incident diabetes diagnosis over seven years since ART initiation when comparing the intervention arm to the control arm. **Panel A** shows the forecasted relative risk of incident diabetes in the intervention (vs. control) arm in the five years following the end of the two-year BMI maintenance intervention. **Panel B** shows the forecasted number of incident diabetes diagnoses averted (per 1,000 person-years) in the intervention (vs. control) arm. **Panel C** shows the forecasted number needed to treat (i.e., the median number of individuals who must experience the BMI maintenance outcome to avert 1 incident diabetes diagnoses). **Panel D** shows the forecasted total number of incident diabetes diagnoses averted in the intervention (vs. control) arm. Across all panels, the median and interquartile range are represented in the box plots, with whiskers extending to 95 percentile range among 1,000 random simulations under each arm. Forecasted population-level outcomes, including the median and 95% uncertainty range, are shown at the bottom each panel. Abbreviations: PWH: people with HIV, ART: Antiretroviral treatment, BMI: Body Mass Index, Black: Black/African American, Het: Heterosexual, WWID: Women Who Inject Drugs, MWID: Men Who inject Drugs, MSM: Men who have Sex with Men, NNT: Number needed to treat to avert one new diabetes diagnosis. NNT is calculated as 1 over absolute reduction in risk of new diabetes diagnosis in the intervention arm compared to control arm.

Relative to the risk of incident diabetes diagnoses in absence of the intervention, Hispanic heterosexual men experienced the largest relative reductions in risk (31% [18–42%]) followed by Black MWID (27% [14–39%]) (**Figure 3B**). White and Hispanic WWID experienced the largest absolute reductions in risk (**Figure 3A**), and the smallest number needed to treat to avert one new diabetes diagnosis (mean NNT=21 persons, **Figure 3C**). At a population level, the greatest number of diabetes diagnoses averted was forecasted among Black MSM (376 [207–555]), followed by Black heterosexual women (341 [212– 476]) during the seven years after ART initiation.

### Differential Impact with Regard to Individuals’ Age and BMI at ART Initiation

The forecasted impact of interventions on diabetes risk was sensitive to individuals’ age at ART initiation. The relative reduction in diabetes risk increased with age, ranging from 15% [6.0–23%] among individuals younger than 20 years to 19% [3–32%] among those aged 60-69 at ART initiation (**Figure 4A-B**). Largest reductions, 21% [-63–63%], were forecasted for individuals aged 70 and older at ART initiation, however, these forecasts carry larger uncertainty due to the smaller population size in this age group.

**Figure 4.**
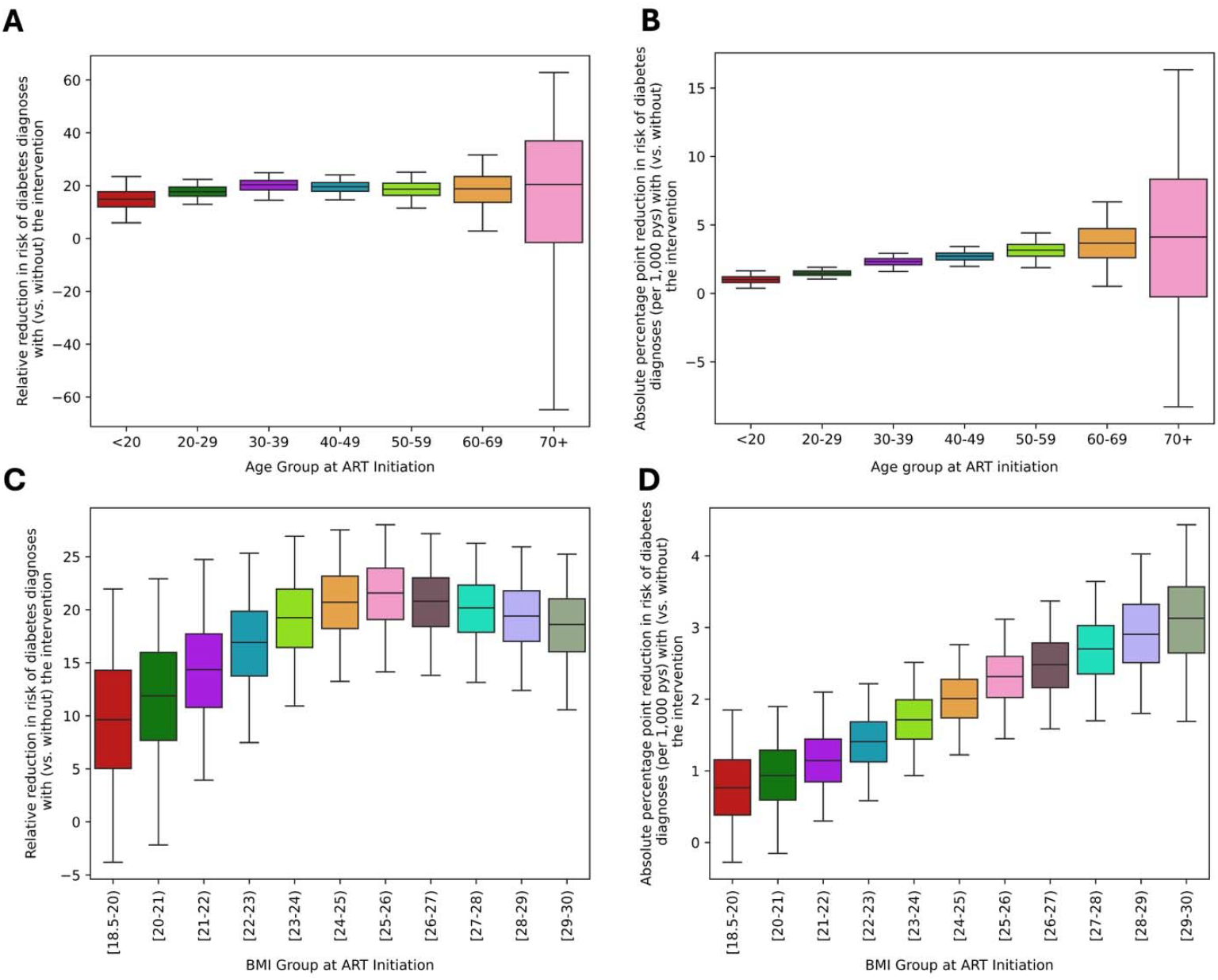
Differential impact of a BMI maintenance intervention at ART initiation on the risk of incident diabetes diagnosis by age and BMI at ART initiation. The forecasted outcomes are compared based on age at ART initiation **(Panels A** and **B**) and BMI ranges at ART initiation (**Panels C** and **D**). **Panels A** and **C** show the forecasted relative reduction in the seven-year risk of diabetes in the BMI maintenance vs. control arms. **Panels B** and **D** show the forecasted absolute percentage point reduction in seven-year risk of diabetes under the intervention arm compared to the control arm. Box plots represent the median and interquartile range of these estimates, with whiskers extending to show the 95 percentile range among 1,000 random simulations under each arm. Abbreviations: ART: Antiretroviral treatment, BMI: Body Mass Index, pys: person-years The large uncertainty ranges observed in the 70+ age group are likely driven by the smaller population size of this age group at ART initiation. For age groups in 20-29, 30-39, 50-59, 70+ and BMI groups in 22-23, 23-24, 24-25, 27-28, 29-30, the outliers generating from simulation randomness are removed from above figures.

Similarly, the intervention’s impact on diabetes risk increased with individuals’ BMI at ART initiation. The absolute reduction in risk over seven years of follow up grew linearly with baseline BMI, ranging from 0.8 [-0.3–1.8] fewer cases per 1000 person-year among individuals starting ART with the lowest BMI (18.5– 19 kg/m^2^) to 3.1 [1.7–4.4] fewer cases per 1000 person-year for those with the highest BMI (28–29 kg/m^2^) (**Figure 4C-D**). The relative reductions in risk also scaled linearly, beginning with a 9.6% [-3.8–22%] reduction among those starting ART with a BMI of 18.5–19 kg/m^2^, and increasing to 22% [14–28%] among individuals starting with a BMI of 25–26 kg/m^2^, after which the reductions stabilized and showed no further improvement for higher BMI categories.

### Sensitivity Analysis

As the intervention’s effectiveness decreased, projected reductions in diabetes risk also decreased, with the number needed to treat rising from 71 cases at 100% effectiveness to 95 at 75%. At the population level, reduced coverage or effectiveness to 75% each, led to a 25% decrease in cases averted, while a 44% reduction in impact occurred when both were reduced simultaneously. Furthermore, subgroups with higher baseline risk (e.g., White and Hispanic WWID) saw a larger reduction in diabetes diagnoses when effectiveness dropped. Subgroups with more new diagnoses (e.g., Black MSM and Black heterosexual women) experienced a greater decrease in diabetes diagnoses when both coverage and effectiveness were reduced, though relative reductions remained consistent across groups (**Supplement S3.1**).

In the one-way sensitivity analysis, the forecasted intervention’s impact was most sensitive to changes in BMI after ART initiation. When BMI increased by 20%, the forecasted reduction in diabetes risk increased by over 300%. Conversely, a 20% reduction in BMI after ART initiation nearly eliminated the intervention’s impact. At the subgroup level, the largest impact was observed among White and Hispanic MSM (**Supplement S3.2**).

## Discussion

Our findings underscore the potential impact of BMI maintenance interventions during the 2 years following ART initiation in reducing diabetes risk among PWH. These findings provide a strong rationale for future clinical trials focused on weight maintenance in the early ART period. The groups likely to benefit most include people who inject drugs, women, and people of color, with Black MSM representing a substantial portion of averted cases, underscoring a critical public health priority. This variability in impact underscores the real-world heterogeneity that can arise, even with perfect coverage and treatment adherence. It emphasizes the necessity of designing interventions that target the highest-risk groups and the importance of tailoring strategies to effectively address the diverse populations aging with HIV in the US.

Previous research indicates a significant link between ART regimens and early weight changes in PWH. INSTIs, recommended as first-line treatments in 2014, became the preferred regimen by 2017 due to their favorable tolerability profile. However, INSTI-based regimens and TAF, have been associated with greater weight gain among ART-naïve individuals compared to older ART classes,^32-34^ and this weight gain disproportionately affects aging women and Black and Hispanic individuals.^18,34,35^ INSTIs may also increase diabetes risk through mechanisms that may be partially but not completely mediated by weight gain.^18,36,37^ The PEARL model uses NA-ACCORD data from 2009-2017, and may underestimate weight gain and diabetes risk due to limited INSTI and TAF use during this period. To the extent that diabetes risk from newer regimens is mediated by weight gain, our forecasted reductions in diabetes risk from the BMI maintenance intervention may underestimate its true impact. Future studies should examine the impact of ART regimen class on weight gain and diabetes, particularly in high-risk subgroups.

Despite the health consequences of obesity and weight gain among PWH, few weight loss programs have been specifically assessed in this population.^38^ Most interventions for PWH have primarily addressed wasting (pre-ART era),^10,15,39^ dyslipidemia (early ART era),^40-42^ or cardiovascular disease risk factors (modern era).^43,44^ Limited studies on weight loss suggest that moderate diet-induced weight loss improves insulin sensitivity in women with HIV, however, PWH take longer to achieve similar results and experience greater decline in fat-free mass.^38,45^ A 12-week randomized controlled trial showed that PWH in an online behavioral weight loss program lost more weight than education-only controls (mean, 4.4± 5.4 kg vs 1.0±3.3 kg; P = 0.02).^46^ However, only 30% achieved a clinically significant (5%) weight loss. These findings underscore the potential of lifestyle interventions for weight management in PWH but emphasizes the need for further research to optimize strategies for this population.

GLP-1 receptor agonists, such as, semaglutide, offer a promising avenue for managing weight among PWH.^47,48^ Initially developed for diabetes, these medications have proven effective in promoting weight loss, improving metabolic health, and addressing wide range of comorbidities.^49-52^ Current clinical guidelines among PWH focus their use among individuals with obesity or prediabetes/diabetes,^53^ but challenges include the difficulty of maintaining long-term weight loss without ongoing treatment and concerns about weight regain if therapy is discontinued. Access to GLP-1 medications in the US remains uneven, as they are often not reimbursed by Medicare or most state Medicaid programs for weight loss alone.^54^ Disparities in access persist even among those with diabetes.^55^ Guidelines also exclude individuals starting ART at normal or overweight levels, as the simultaneous initiation of GLP-1 medications and ART is complicated by limited evidence, potential drug-drug interactions, and side effects that could impact ART adherence. Moreover, GLP-1 has been linked to muscle mass loss, posing long-term challenges for insulin regulation and physical function, particularly in older PWH.^47^ Further research is critical to evaluate the safety, effectiveness, and address access inequities among PWH, allowing for more personalized and effective weight management strategies.

Our findings are subject to several limitations. NA-ACCORD data on weight and BMI are collected as part of routine care, not as prospectively collected with regular frequency and uniform calibration except in interval cohorts. To ensure model reliability, we focused on BMI measurements at ART initiation and two years after initiations, as these provided the highest quality data and are key milestones for comorbidity risk. However, this approach excludes later changes in BMI and impact on comorbidity risk. Future studies should address gradual weight changes and long-term program effects. Additionally, while useful as a screening tool, BMI is not a direct measure of body fat and not an accurate measure to diagnose overweight or obesity.^56^ BMI overestimates body fat in athletes with high muscle mass and in patients with edema, while underestimating it in sarcopenic individuals. It also does not account for fat distribution, which is important for metabolic risk.^57^ Other measurements, such as waist circumference and Dual-Energy X-ray Absorptiometry (DEXA), may offer more insight but are rarely used in PWH. Finally, ART regimens were excluded from this analysis. Further research is needed to clarify ART drug interactions and their effects on weight and comorbidities in PWH.

Despite these limitations, high-fidelity simulation models like PEARL provide valuable insights into novel interventions. Our findings lay the groundwork for future clinical trials and highlight key data gaps, while identifying populations that would benefit most from this intervention to inform recruitment strategies. Sensitivity analysis underscores the achievable impact among populations facing barriers to coverage and adherence. Finally, PEARL’s strength lies in leveraging NA-ACCORD data to inform individual-level risk functions for comorbidities, representing 15 distinct subgroups of PWH. Understanding the complex interplay between various factors—such as BMI, comorbidities, adherence, and population heterogeneity—will be essential in guiding clinical trial design and shaping policies to better address the needs of diverse PWH populations.

Our findings suggest that maintaining weight during the first two years after ART initiation holds promise for improving long-term health outcomes. However, the observed heterogeneity in outcomes underscores the importance of caution. BMI maintenance interventions must be tailored to the unique needs of specific subgroups within the population to ensure effectiveness.

## Author Contributions

PK and KA conceived the study. LH and KA provided data from NA-ACORD. PK, YZ, and KR conducted the simulation analysis. KE, KG, KL, and KK provided clinical and statistical feedback. YZ, EH, KR and PK developed the methodological supplement. PK and KA wrote the manuscript. KL, KK, KE, KG, PR, JK, KB reviewed and revised the manuscript. All authors reviewed the results and manuscript and approved the final version.

## Data availability

PEARL source code is available on GitHub (https://github.com/PearlHivModelingTeam/pearlModel) and under MIT License.

